# Distinct response of resting-state brain networks to psilocybin in autism

**DOI:** 10.64898/2026.08.19.26360794

**Authors:** Tobias P. Whelan, Mihail Dimitrov, Lucas G.S. França, Claire L. Ellis, Francesca Moruzzi, Francesca M. Ponteduro, Johanna Kangas, Nermin Khalil, Yan Ge, Naoise Mulcrone, Glynis Ivin, Dafnis Batallé, Eileen Daly, Ekaterina Malievskaia, Nicolaas A. Puts, Declan G.M. Murphy, Gráinne M. McAlonan

**Affiliations:** Department of Forensic and Neurodevelopmental Science, Institute of Psychiatry, Psychology & Neuroscience, King’s College London, London, United Kingdom; N1 Bio Corp., Miami, Florida, USA; Department of Early Life Imaging, School of Biomedical Engineering & Imaging Sciences, King’s College London, London, United Kingdom; School of Computer Science, Faculty of Science and Environment, Northumbria University, Newcastle upon Tyne, United Kingdom; South London and Maudsley NHS Foundation Trust Pharmacy, London, United Kingdom; NIHR Biomedical Research Centre: Maudsley, South London and Maudsley NHS Foundation Trust and King’s College London, London, United Kingdom; MRC Centre for Neurodevelopmental Disorders, King’s College London, London, United Kingdom

**Author notes:** **Corresponding author:** Grainne McAlonan, Department of Forensic and Neurodevelopmental Sciences, Institute of Psychiatry, Psychology & Neuroscience, King’s College London, UK.

## Abstract

**Importance:** There is increasing interest in the potential of psilocybin to treat mental health and neurodevelopmental conditions. At high doses, the therapeutic benefit of psilocybin is linked to greater functional connectivity or integration between large-scale brain networks which underpin mood, emotion and cognition. However, it is unknown how the brain responds to psilocybin in autism – a condition characterised by both altered functional connectivity and differential response to drugs. Thus, a first step before clinical trials of psilocybin involving autistic people, is to evaluate the response of the autistic brain to psilocybin, initially at low dose.

**Objective:** Low doses of psilocybin were used to test the hypothesis that the functional connectivity of large-scale resting-state brain networks respond differently in autistic and non-autistic adults.

**Design:** The ‘PSILAUT’ study had a pseudo-randomised, cross-over, double-blind, case-control design. There was no evaluation of clinical efficacy. PSILAUT was not a Clinical Trial according to UK regulations. Data collection was conducted from January 2023 to August 2024.

**Setting:** Single-centre, study conducted at the Institute of Psychiatry, Psychology & Neuroscience, King’s College London, London, United Kingdom.

**Participants:** Adult (> 18 years) participants with and without an autism spectrum disorder (ASD) diagnosis were recruited and matched for age, sex and IQ. Autistic participants were included if they had an existing diagnosis (DSM-IV, DSM-5 or ICD-10 criteria).

**Exposures:** A single oral dose of 2 or 5 mg psilocybin or (inactive) placebo administered on separate visits at least one week apart.

**Main Outcomes and Measures:** Resting-state fMRI was acquired to investigate the change in functional connectivity within and between brain networks, as measures of network integrity and integration, respectively.

**Results:** A total of 67 participants were recruited (18-58 years at first visit; 30 non-autistic participants, mean [SD] age, 30.0 [8.3] years, 15 males [50%] and 37 autistic participants, mean [SD] age, 28.6 [9.2] years, 19 males [51%]).

We report for the first time that the autistic brain responds differently to low doses of psilocybin, despite no group differences in network connectivity in the baseline placebo condition. 5 mg psilocybin elicited the greatest shifts in functional connectivity in both groups, but in different directions. In non-autistic participants only, on average, after 5 mg psilocybin within-network connectivity of the frontoparietal (β = −0.053, T = −2.73, FDR-corrected *P* value = 0.027, Cohen *d* = −0.71) and limbic networks (β = −0.087, T = −2.59, FDR-corrected *P* value = 0.021, Cohen *d* = −0.61) decreased. In contrast, in autistic participants, 5 mg psilocybin increased between-network connectivity (i.e. integration) of higher-order and attentional networks, but decreased connectivity between the same networks in non-autistic participants (default mode and frontoparietal networks, dose × group interaction: β = 0.053, T = 2.91, FDR-corrected *P* value = 0.042; dorsal and ventral attention networks, dose × group interaction: β = 0.059, T = 2.31, FDR-corrected *P* value = 0.015). Across the whole sample, the extent to which psilocybin elicited an increase in connectivity between higher-order (β = 0.33, T = 2.16, FDR-corrected *P* value = 0.036) and attentional (β = 0.36, T = 2.43, FDR-corrected *P* value = 0.036) networks was positively correlated with core autistic traits quantified using the Autism Quotient.

**Conclusions and Relevance:** Functional brain networks that support mood, emotion and cognition are more responsive to low dose psilocybin in autistic adults compared to non-autistic adults. Given that increased network integration is associated with clinical utility, future applications of psilocybin in autistic people should include the evaluation of low doses.

**Key Points:** *Question:* Adult autism is commonly complicated by co-occurring mental health difficulties. Psilocybin is showing promise across psychiatry but we do not know whether the autistic brain responds differently to psilocybin.

*Findings:* 5 mg psilocybin increased between-network connectivity (or integration) in autistic participants only; a pattern usually seen after higher doses in non-autistic people and linked to therapeutic benefit in other neuropsychiatric conditions.

*Meaning:* Future applications of psilocybin in autistic people should prioritise evaluation of lower doses.

## Introduction

The classic psychedelic psilocybin acts on the serotonin system and is increasingly showing promise across psychiatry, with clinical utility in mental health conditions which are common in autistic people^1,2^. We have previously reported that the pharmacological *responsivity* of the resting-state brain networks which underpin social cognition, emotion, behaviour and mental health is different in non-autistic and autistic people^3^. This includes the serotonin system, long implicated in the autistic phenotype and the target of several psychiatric medications^4^. However, we do not know if the autistic brain responds differently specifically to psilocybin. This is important to establish as psilocybin increasingly becomes available in mental health settings globally.

In non-autistic people, psilocybin at high doses (e.g. 25 mg) increases the functional connectivity between resting-state networks^5–8^ and this action on network integration has been linked to improved mental health^9^. However, because our prior work found both greater and atypical neural responses to serotonergic pharmacological challenge in autistic compared to non-autistic people^10^, we elected to use low oral doses of psilocybin active at the 5HT_2A_ receptor^11^ (reported to have lower availability in autism^12^). In addition, low doses meant we could minimize marked (and potentially distressing^13^) psychedelic experiences which could confound interpretation. The study could also be conducted without specialised psychological support (for example^14^), given the adjustments needed for autism are not known.

Hence, resting-state brain fMRI data acquired from autistic and non-autistic adults was compared following a double-blind single low oral dose of 2 or 5 mg psilocybin or placebo in a pseudo-randomised order across three separate visits (see Whelan et al.^15^ for a description of the full ‘PSILAUT’ protocol). Subjective effects were screened using the gold-standard 5-dimensional altered states of consciousness questionnaire (5D-ASC)^16^. Recognising that autistic traits may exist as a continuum in the population^17^, we also explored any correlations of psilocybin-elicited network connectivity alterations with self-reported core autistic traits (measured by the autism quotient (AQ)^18^) across the whole cohort.

We held *a priori* hypotheses: i) Based on evidence that higher doses of psilocybin reduce within-network and increase between-network connectivity^6,9,19^, we expected that this pattern might start to emerge at 5 mg psilocybin. ii) Based on our prior work showing that pharmacological (including serotonergic) challenge primarily increases between-network connectivity in autistic adults^10^, we predicted that psilocybin would preferentially increase between-network connectivity in autistic compared to non-autistic participants.

## Methods & Materials

### Study procedures

The study was an Investigator-Initiated Study sponsored by King’s College London and co-Sponsored by South London and Maudsley NHS Foundation Trust (SLaM). Written informed consent was obtained from all participants in accordance with the Helsinki Declaration of 1964, as revised in 2013 and the study procedures were approved by Dulwich Research Ethics Committee (21/LO/0795). Psilocybin was stored in our on-site Maudsley Hospital pharmacy and administered in our study under appropriate licences from the UK Home Office. Our study did not address safety or clinical efficacy, and the UK Medicines and Health Regulatory Authority (MHRA) confirmed that our protocol was therefore not a clinical trial of an Investigational Medicinal Product (IMP) as defined by the EU Directive 2001/20/EC. For transparency, our study was pre-registered on ClinicalTrials.gov (NCT05651126). The study was part funded by Compass Pathfinder Ltd with infrastructure support from the NIHR-Maudsley Biomedical Research Centre at South London and Maudsley NHS Foundation Trust and King’s College London. Psilocybin was supplied at no cost to the investigating team as COMP360 psilocybin, a proprietary synthetic formulation of crystalline psilocybin owned by Compass Pathways plc, London, United Kingdom.

The complete ‘PSILAUT’ study protocol procedures are described in detail elsewhere (please see Whelan et al.^15^). A single dose of 2 mg, 5 mg psilocybin or (inactive) placebo was administered on three separate visit days, at least 6 days apart. The 2 mg dose preceded the 5 mg dose as approved by the ethics committee. Thus, any adverse response in initial study visits, for example an increase in blood pressure, would permit unblinding by the Chief Investigator to avoid exposing that individual to a higher dose of psilocybin.

### Participants

Participants were screened by an experienced nurse or doctor prior to enrolment and an in-person health check including blood pressure measurement was conducted by a nurse or doctor at the beginning of the visit, following drug administration and at the end of every visit. This clinical cover was available throughout the duration of the visit. A clinical autism diagnosis from a recognised UK assessment service was accepted. Participants with syndromic autism (of known genetic cause e.g. Fragile X syndrome) were excluded. Other inclusion criteria included the ability to provide informed consent; an IQ > 70; being over 18 years old; no co-occurring psychiatric illness such as major mood disorder or psychotic illness; no physical illness such as high blood pressure and no history of seizures or diagnosis of epilepsy. People taking medications that affect serotonin (such as selective serotonin-reuptake inhibitors) were excluded. Given that autistic people have high rates of ADHD, those taking stimulants were eligible and they were asked to omit their medication on the day of testing. On each visit, participants were screened for illicit substances and pregnancy (where appropriate) by providing a urine sample.

A total of 67 individuals completed the study, including n = 30 non-autistic and n = 37 autistic participants. The non-autistic and autistic group were well-balanced for sex, age and IQ as none of these metrics were significantly different between groups. Demographic information is provided in **Table 1** below.

**Table 1.**
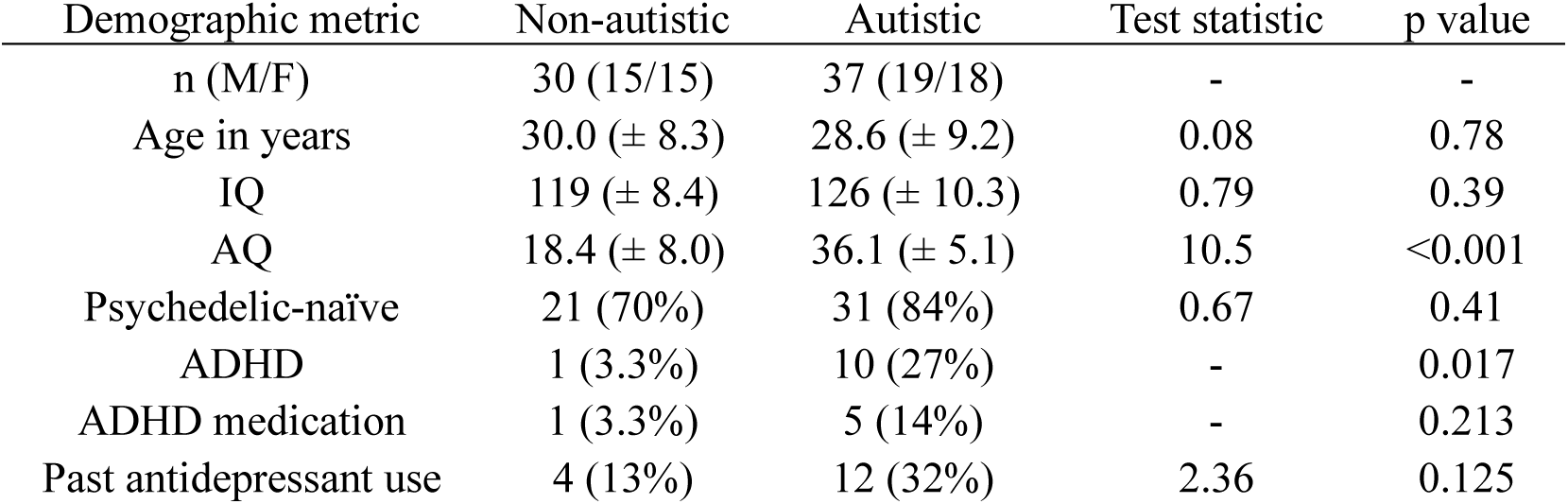
Demographic information for all participants in the ‘PSILAUT’ study. Group differences were assessed and test statistics (either F or X^2^ where appropriate) and p values obtained using the non-parametric Brown-Forsythe, and Chi-squared or Fisher’s Exact test for categorical variables. Where appropriate, standard deviations (for age, IQ and AQ) or percentage proportions are provided in brackets. n = 1 trans autistic participant (assigned male at birth). ‘Psychedelic-naïve’ was defined as no previous lifetime use of a classic psychedelic (e.g. psilocybin (-containing mushrooms), LSD, DMT). ‘ADHD’ (Attention Deficit Hyperactivity Disorder) refers to formal (self-reported during the screening process) diagnoses of ADHD. ‘Past antidepressant use’ refers to prior lifetime use of serotonergic antidepressant medications such as SSRI/SNRIs and/or tricyclic antidepressants. ‘ADHD medication’ refers to stimulant ADHD medications such as lisdexamfetamine or Concerta, all participants abstained from taking ADHD medications on study visit days. Concurrent medications (including details of ADHD medications) of study participants not included here are provided in **Supplementary Table 1**. ADHD, attention-deficit hyperactivity disorder; AQ, autism quotient; F, female; IQ, intelligence quotient (as measured using the Wechsler Abbreviated Scale of Intelligence Second Edition, WASI-II^20^ (two-scale)); M, male; SSRI, selective-serotonin reuptake inhibitor; SNRI, serotonin-norepinephrine reuptake inhibitor.

| Demographic metric | Non-autistic | Autistic | Test statistic | p value |
| --- | --- | --- | --- | --- |
| n (M/F) | 30 (15/15) | 37 (19/18) | - | - |
| Age in years | 30.0 (± 8.3) | 28.6 (± 9.2) | 0.08 | 0.78 |
| IQ | 119 (± 8.4) | 126 (± 10.3) | 0.79 | 0.39 |
| AQ | 18.4 (± 8.0) | 36.1 (± 5.1) | 10.5 | <0.001 |
| Psychedelic-naïve | 21 (70%) | 31 (84%) | 0.67 | 0.41 |
| ADHD | 1 (3.3%) | 10 (27%) | - | 0.017 |
| ADHD medication | 1 (3.3%) | 5 (14%) | - | 0.213 |
| Past antidepressant use | 4 (13%) | 12 (32%) | 2.36 | 0.125 |

### Baseline questionnaires

#### Autism quotient (AQ)

AQ scores were collected before the first visit as part of a larger baseline characterisation online using PsyTools (Delosis Ltd., London). The AQ is a validated self-administered 50-item questionnaire that quantifies core autistic traits^18^. Four non-autistic and 3 autistic participants did not complete the AQ.

#### MRI data acquisition

Structural and functional MRI data were acquired on a General Electric SIGNA^TM^ Premier 3T MRI scanner at the Centre for Neuroimaging Sciences, King’s College London, London, United Kingdom. A high resolution T1-weighted anatomical scan was acquired each visit. Structural scans used an MPRAGE PROMO sequence with the following parameters: voxel size = 1 mm^3^ isotropic, flip angle (FA) = 8°, inversion time (TI) = 860 ms, field of view (FOV) = 256 mm; matrix = 256 x 256. A functional scan was acquired during resting-state with the participant asked to keep their eyes open and fixated on a cross. The resting-state scan used a single-echo EPI sequence with the following parameters: voxel size = 2.70 mm^3^ isotropic; multiband factor 4; TR = 0.933s; 525 total volumes (515 used in final analyses); TE, 32 ms; 52 interleaved slices; 2.70 mm slice thickness; 2.70 mm^3^ isotropic voxel size; FOV, 221 mm; FA, 60°. Resting-state MRI data were collected an average of 77 (± 8.9) mins post-administration of psilocybin or placebo in non-autistic participants and 75 (± 7.0) mins post-administration in autistic participants, which is expected to be during peak drug effects at the low doses of psilocybin used^11^.

#### Structural MRI data preprocessing

Structural MRI images were manually inspected to ensure sufficient signal-to-noise ratio and data quality. Preprocessing of both structural and functional MRI data was performed using *fMRIPrep* 22.0.2^21^. The single T1-weighted anatomical image was skull-stripped and segmented into cerebrospinal fluid (CSF), white matter and grey matter. Brain surfaces were reconstructed using FreeSurfer 7.2.0.^22^ and normalised to standard Montreal Neurological Institute (MNI) space (MNI152NLin6Asym).

#### Functional MRI data preprocessing

Functional MRI images were manually inspected to ensure sufficient data quality, this included identification of artifacts such as warping, ghosting, distortions or blurring. Preprocessing of functional MRI data was also performed using *fMRIPrep* 22.0.2^21^. The BOLD fMRI images were slice time corrected and then realigned to their native space by applying the transforms to correct for head motion. A BOLD reference volume was co-registered to the respective structural T1 reference image using FreeSurfer 7.2.0.^22^. Probabilistic masks for white matter and CSF were generated in anatomical space to allow for component-based noise correction at a later stage of processing (CompCor^23^). Automatic removal of motion artifacts (noise regression) using independent component analysis (ICA-AROMA^24^) was performed on the preprocessed BOLD timeseries in MNI space after removal of non-steady state volumes and spatial smoothing with a Gaussian kernel of 6 mm full-width half-maximum (FWHM). Average mean framewise displacement (mean FD) was higher in autistic participants (placebo, 0.14 (± 0.07); 2 mg, 0.16 (± 0.10) & 5 mg, 0.16 (± 0.07)) compared to non-autistic participants (placebo, 0.10 (± 0.03); 2 mg, 0.11 (± 0.03) & 5 mg, 0.13 (± 0.07)) across all conditions, however, there were no significant group differences at each dose. This suggests that autistic participants have inherently increased motion in the scanner (as reported previously^25^) rather than a psilocybin-specific effect. No scans were above the mean FD exclusion threshold (> 0.5 mm^26^). Further preprocessing and denoising was conducted in CONN v21.a^27^ which included the removal of the first 10 (non-steady state) volumes, regression of white matter and CSF noise components, high-pass filtering above 0.008 Hz and linear detrending.

#### Functional network analysis

Preprocessed BOLD timeseries from each scan in MNI space were parcellated into 100 cortical brain regions of interest (ROIs) according to the Schaefer atlas^28^. Pearson’s correlation was used to compute the linear association between the average timeseries of each pair of ROIs as a measure of functional brain connectivity. These values were then Fisher-transformed into z values (Fisher-transformed correlation coefficients). Each node (of 100 total according to the Schaefer atlas) was assigned to a resting-state brain network according to the Yeo-7 Network atlas^29^: default mode (DMN); frontoparietal (FPN); ventral attention (or salience) (VAN); dorsal attention (DAN); limbic (LN); somatomotor (SMN) & visual (VN) networks. Within-network connectivity was defined as the mean connectivity between each ROI that comprised a given Yeo-7 Network. Between-network connectivity was defined as the mean connectivity between ROIs that comprised two different Yeo-7 Networks. This analytical framework has been applied previously to identify response differences in functional networks following drug challenge in autistic and non-autistic people^10^.

#### Subjective effects

The ‘gold standard’ 5D-ASC questionnaire^16^ was used at the end of the participant visit approximately 4 hours post-administration (non-autistic: 235 (± 25.4) mins and autistic: 227 (± 30.1) mins) to quantify the extent of alterations in waking consciousness (or ‘subjective effects’). This 94-item self-administered questionnaire generates a total score from five dimensions: oceanic boundlessness (OB, 27 items); anxious ego dissolution (AED, 21 items); visionary restructuralisation (VR, 18 items); auditory alterations (AA, 15 items) and reduction of vigilance (RoV, 12 items), each item scored from 0 to 10. One autistic participant’s questionnaire (5 mg session) was incomplete and was excluded from analyses.

### Statistical analyses

#### Within-network & between-network functional connectivity

We adopted an exploratory approach in which an omnibus linear mixed effects (LME) model was fit using the lme4 package in R Studio 2022.07.0^30^ given by the expression:

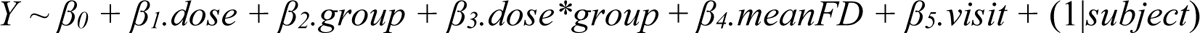

*Y* denotes the within- or between-network connectivity, *dose* (0, 2 or 5 mg) and *group* (non-autistic or autistic) were fixed effects, mean framewise displacement (*meanFD*) was included as a covariate to account for head motion^26^, *visit* is the study visit order, and *subject* accounted for the random effect.

First, dose × group interactions tested whether each dose of psilocybin altered within- and between-network connectivity and whether effects differed by group, and the main effect of group tested for baseline (i.e. placebo) group differences in within- and between-network connectivity; p values for fixed effects were obtained using a two-sided permutation test of 10,000 repetitions and adjusted for multiple comparisons using the Benjamini-Hochberg false discovery rate (FDR) method^31^.

Post-hoc analyses of the main effect of dose were conducted for each group separately where the omnibus model yielded a significant dose effect or dose × group interaction (before correction). Post-hoc pairwise dose comparisons were adjusted for multiple comparisons using FDR. Effect size (Cohen’s *d*) was calculated for each psilocybin dose relative to placebo to quantify the magnitude of connectivity change.

Total between-network connectivity of each network is reported as the sum of change in connectivity of the network with all the between-network pairs (as previously described^10^) and the overall change in between-network connectivity of each group was compared at each psilocybin dose using unpaired t-tests (after normality was assessed with the Shapiro-Wilk test, p > 0.05).

Age and sex were not included as covariates in the main analyses because they did not have significant main effects after FDR correction when included in the omnibus model.

#### Subjective effects

In an extended analysis, the effect of psilocybin on overall subjective effect intensity (5D-ASC total score) and specific dimensions of the subjective experience (5D-ASC dimension scores) was compared for non-autistic and autistic participants. The statistical approach was as described in the previous section for network connectivity; the LME was given by the expression: *Y* ∼ *β_0_ + β_1_.dose + β_2_.group + β_3_.dose*group + β_4_.visit +* (1|*subject*). As there were no significant group differences in this measure (total and dimension scores) at any dose, this measure was not explored further post-hoc.

#### Relationship of network connectivity changes with autistic traits

Finally, a change score (i.e. Δ5 mg – 0 mg) was calculated for within- and between-network connectivity significantly shifted by psilocybin and a linear regression was fit to establish whether change in network connectivity after psilocybin was associated with the change in the extent of core autistic traits measured by total AQ score across the whole group at baseline; linear regression models were adjusted for motion (mean FD) and visit order, as in the network connectivity analyses.

## Results

### Study cohort

Thirty non-autistic (15 female) and thirty-seven (18 female) autistic individuals were included, and 174 study visits were completed: 61 placebo visits (26 non-autistic and 35 autistic), 61 2 mg visits (27 non-autistic and 34 autistic) and 52 5 mg visits (23 non-autistic and 29 autistic). There was one trans female autistic participant (assigned male at birth). Two non-autistic and two autistic participants had elevated blood pressure after the 2 mg dose and were excluded from the 5 mg dose visit. One non-autistic participant began SSRI treatment following visit one and was excluded from subsequent visits. Twenty-one non-autistic and 29 autistic participants completed all three visits. Visits were at least 6 days apart to ensure complete drug washout.

### Resting-state network connectivity in autistic and non-autistic participants did not differ at baseline

First, baseline (i.e. placebo) group differences in network connectivity were assessed. There was a significant main effect of group for within-network connectivity of the ventral attention network, which was higher in autistic participants at baseline, however, this did not survive correction for multiple comparisons (β = 0.045, T = 1.90, p_unc_ = 0.033, p_FDR_ = 0.186). Thus, there was no robust difference at baseline between the autistic and non-autistic groups in our sample for any network (within- and between-networks) investigated.

### Low dose psilocybin differentially alters network connectivity in autistic and non-autistic people

Next, dose × group interactions and main effects of dose were characterised, establishing the effect of psilocybin dose on within- and between-network connectivity and whether this differed in non-autistic and autistic individuals. Where the LME model yielded significant dose × group interactions and/or main effects of dose before correction, post-hoc testing of the dose effect in each group separately was conducted.

### Within-network connectivity summary

5 mg psilocybin reduced connectivity within the FPN and LN in non-autistic participants but did not shift within-network connectivity in autistic participants.

Specifically, there was a significant dose × group interaction for the DMN after 5 mg psilocybin, but this did not survive correction (β = 0.054, T = 2.25, p_unc_ = 0.027, p_FDR_ = 0.072). There was a significant main effect of 5 mg psilocybin on the FPN (β = −0.058, T = −2.93, p_FDR_ = 0.028) and LN (β = −0.074, T = −2.48, p_FDR_ = 0.041). Post-hoc analyses of the dose effect in each group revealed that 5 mg psilocybin decreased FPN and LN connectivity in non-autistic participants **(Supplementary Figure 1)**; the dose effect did not reach significance (uncorrected) for within DMN connectivity in non-autistic (β = −0.036, T = −1.94, p_unc_ = 0.055) or autistic (β = 0.018, T = 1.16, p_unc_ = 0.25) participants. The magnitude and direction (by effect size) of within-network shifts in connectivity after 2 mg and 5 mg psilocybin is shown for both groups in **Figure 1** and the main results are summarised in **Table 2**.

**Figure 1.**
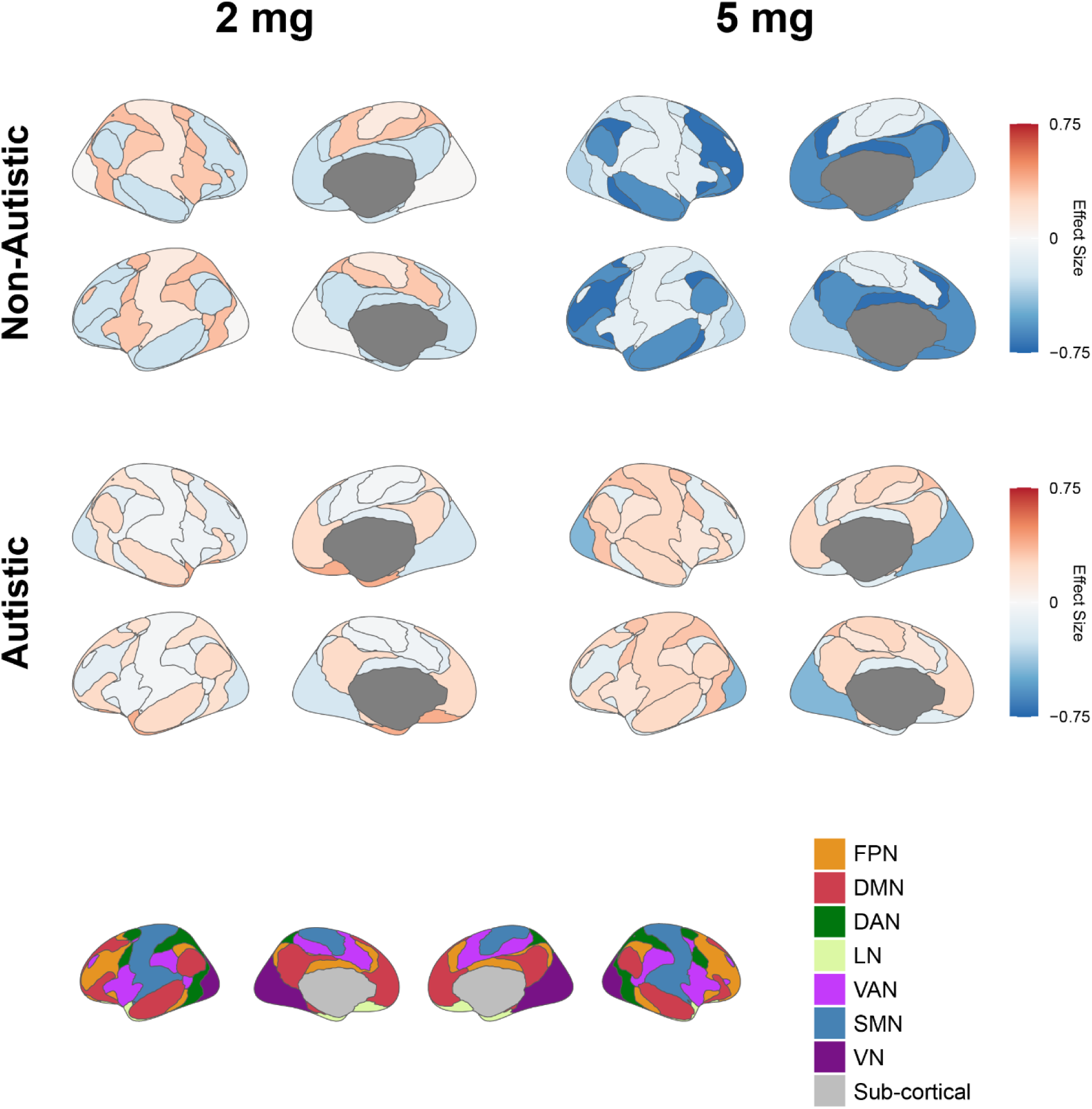
Within-network connectivity alterations in response to 2 and 5 mg psilocybin in non-autistic and autistic participants. Brain plots illustrate the shift in connectivity (represented by Cohen’s d effect size) between placebo and 2 or 5 mg psilocybin. Darker blue regions after 5 mg psilocybin in non-autistic participants represent generally decreased within-network connectivity; the most prominent shifts were in the FPN (d = −0.71) and LN (d = −0.61) which were significantly decreased by 5 mg psilocybin in the non-autistic group. The Yeo-7 network atlas^29^ (bottom) provides a legend key to identify functional networks. DMN, default mode; FPN, frontoparietal; VAN, ventral attention; DAN, dorsal attention; LN, limbic; SMN, somatomotor & VN, visual network.

**Table 2.**
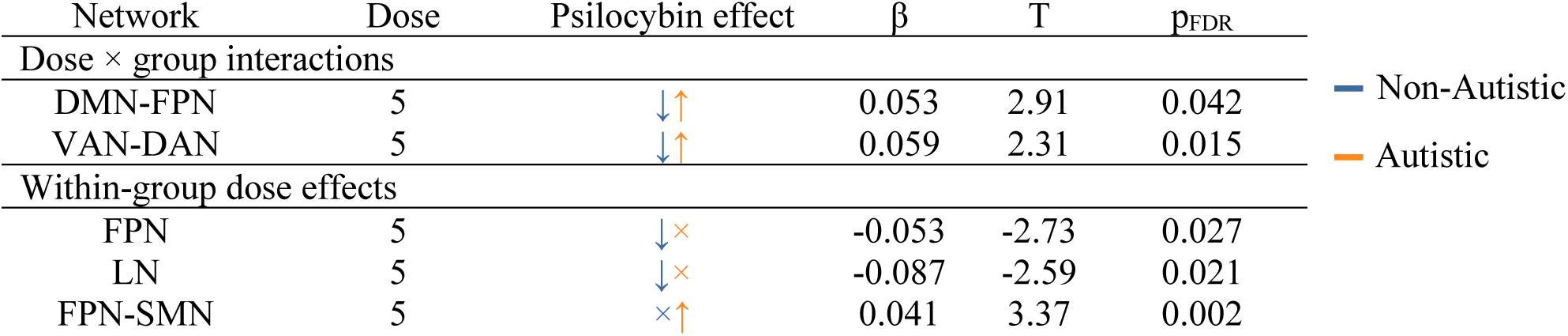
Summary of main findings. All results shown in this table were statistically significant after correction for multiple comparisons (p_FDR_ < 0.05). The blue and orange arrows represent the directionality of the result in the non-autistic and autistic groups, respectively. Adjacent arrows represent dose *×* group interactions. Single arrows represent the within-group main effect of dose. DAN, dorsal attention network; DMN, default mode network; FPN, frontoparietal network; LN, limbic network & VAN, ventral attention network.

### Between-network connectivity summary

5 mg psilocybin decreased between network connectivity of DMN-FPN and VAN-DAN in the non-autistic group, but increased connectivity in these networks in the autistic group; between network FPN-SMN connectivity was also increased in autistic participants.

Specifically, there was a significant dose × group interaction for DMN-FPN and VAN-DAN connectivity after 5 mg psilocybin (**Table 2**); 5 mg psilocybin decreased connectivity in the non-autistic group, but increased connectivity in the autistic group. Post-hoc dose effects examined in each group separately (reported uncorrected) revealed that DMN-FPN connectivity was decreased in non-autistic participants (β = −0.031, T = −2.20, p_unc_ = 0.030), and did not reach significance in autistic participants (β = 0.022, T = 1.86, p_unc_ = 0.066). VAN-DAN connectivity was increased in autistic participants (β = 0.036, T = 2.11, p_unc_ = 0.037) but not non-autistic participants (β = −0.026, T = −1.28, p_unc_ = 0.20).

There was a significant dose × group interaction for FPN-SMN connectivity prior to correction for multiple comparisons (β = 0.041, T = 1.92, p_unc_ = 0.047, p_FDR_ = 0.28), which was increased to a greater extent in autistic participants. Post-hoc analysis revealed a significant main effect of dose for FPN-SMN in the autistic group.

The magnitude (shown by effect size) for change in between-network connectivity after 2 mg and 5 mg psilocybin compared to placebo is shown for non-autistic and autistic participants in **Figure 2** and the main results are summarised in **Table 2**.

**Figure 2.**
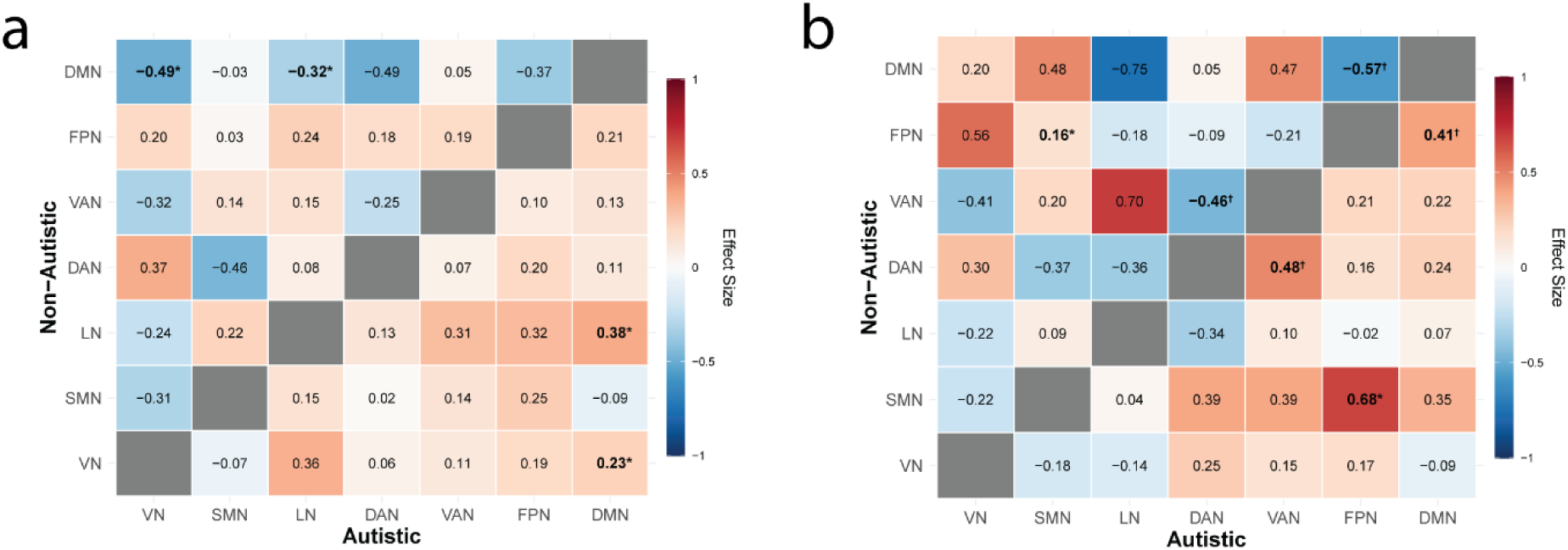
Between-network connectivity alterations in non-autistic and autistic participants at 2 mg and 5 mg psilocybin. Values represent the effect size (Cohen’s d) for **(a)** 2 mg and **(b)** 5 mg psilocybin compared to placebo. Statistically significant dose *×* group interactions prior to correction for multiple comparisons are shown in bold with an asterisk (*p_unc_ < 0.05), and those that survive FDR correction are shown in bold with a dagger (^†^p_FDR_ < 0.05). The 2 mg psilocybin dose predominantly impacted between-network connectivity of the DMN; DMN connectivity with the LN and VN was decreased in non-autistic participants and increased in autistic participants, but the dose *×* group interactions did not survive correction. After 5 mg psilocybin, DMN-FPN and VAN-DAN connectivity was decreased in non-autistic participants and increased in autistic participants. FPN-SMN connectivity was also increased to a greater extent in autistic participants after 5 mg psilocybin but the dose *×* group interaction did not survive correction. DMN, default mode; FPN, frontoparietal; VAN, ventral attention; DAN, dorsal attention; LN, limbic; SMN, somatomotor & VN, visual network.

Despite on average group differences, there was individual variation in responses to psilocybin in both the non-autistic and autistic group. The heterogenous trajectories of change in network connectivity after 5 mg psilocybin compared to placebo for networks with significant dose × group interactions in the main analysis are shown in **Figure 3**.

**Figure 3.**
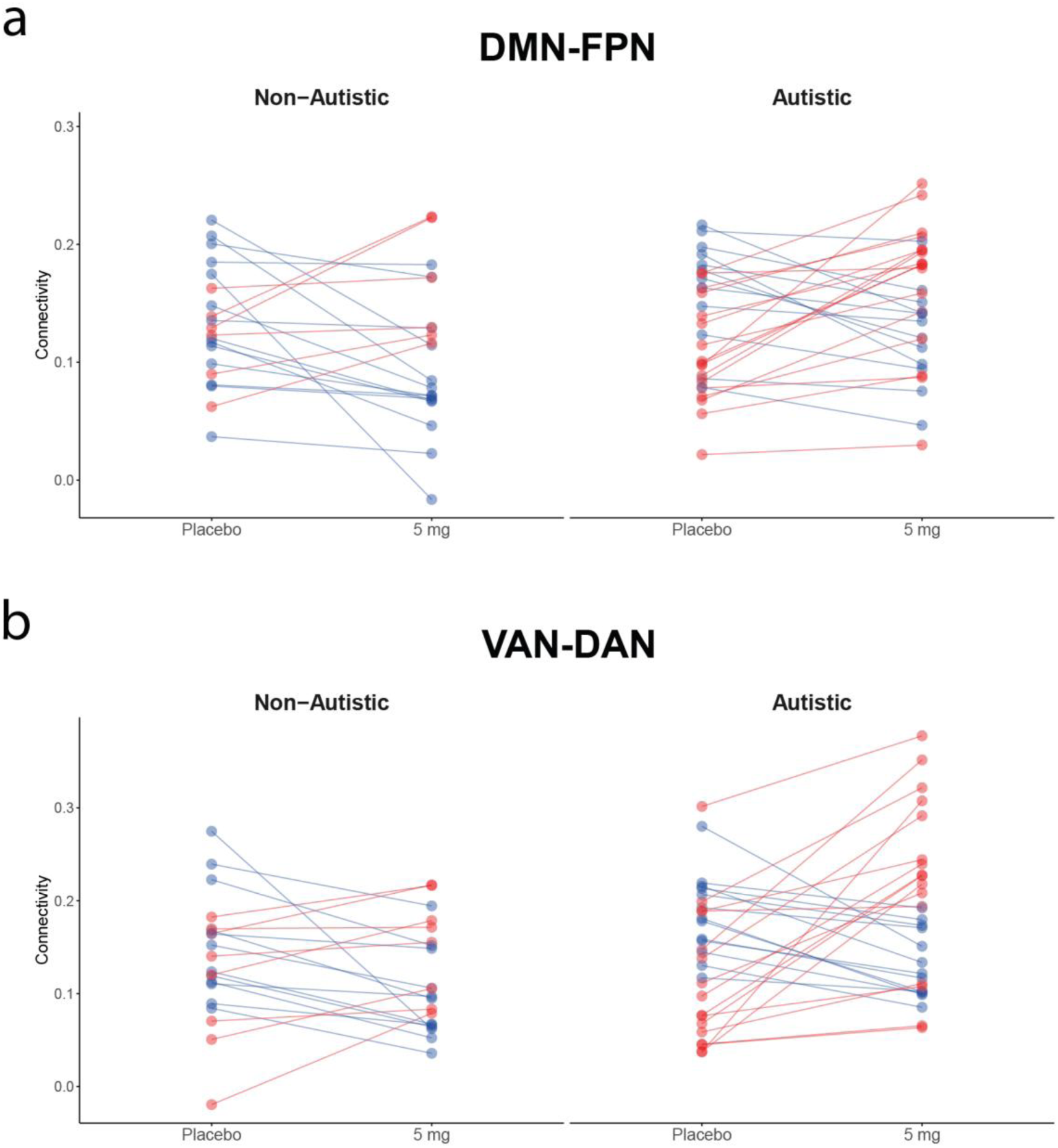
Individual trajectories for the shift in resting-state network connectivity at 5 mg psilocybin in non-autistic and autistic individuals. Individual increases (red) and decreases (blue) in connectivity are shown for between-network connectivity of the **(a)** default mode-frontoparietal network (DMN-FPN) and **(b)** ventral attention-dorsal attention network (VAN-DAN).

### Low dose psilocybin increases overall between-network connectivity to a greater extent in autism

Next, between-network connectivity was aggregated for each network to compare the global impact of psilocybin on between-network connectivity in non-autistic and autistic participants following methods previously reported^10^. Overall, psilocybin shifted between-network connectivity in autistic individuals to a significantly greater extent than non-autistic individuals at both 2 mg (T = 3.34, p_FDR_ = 0.006) and 5 mg (T = 4.42, p_FDR_ = 0.002) psilocybin. Total between-network connectivity for each resting-state network is shown in **Figure 4**.

**Figure 4.**
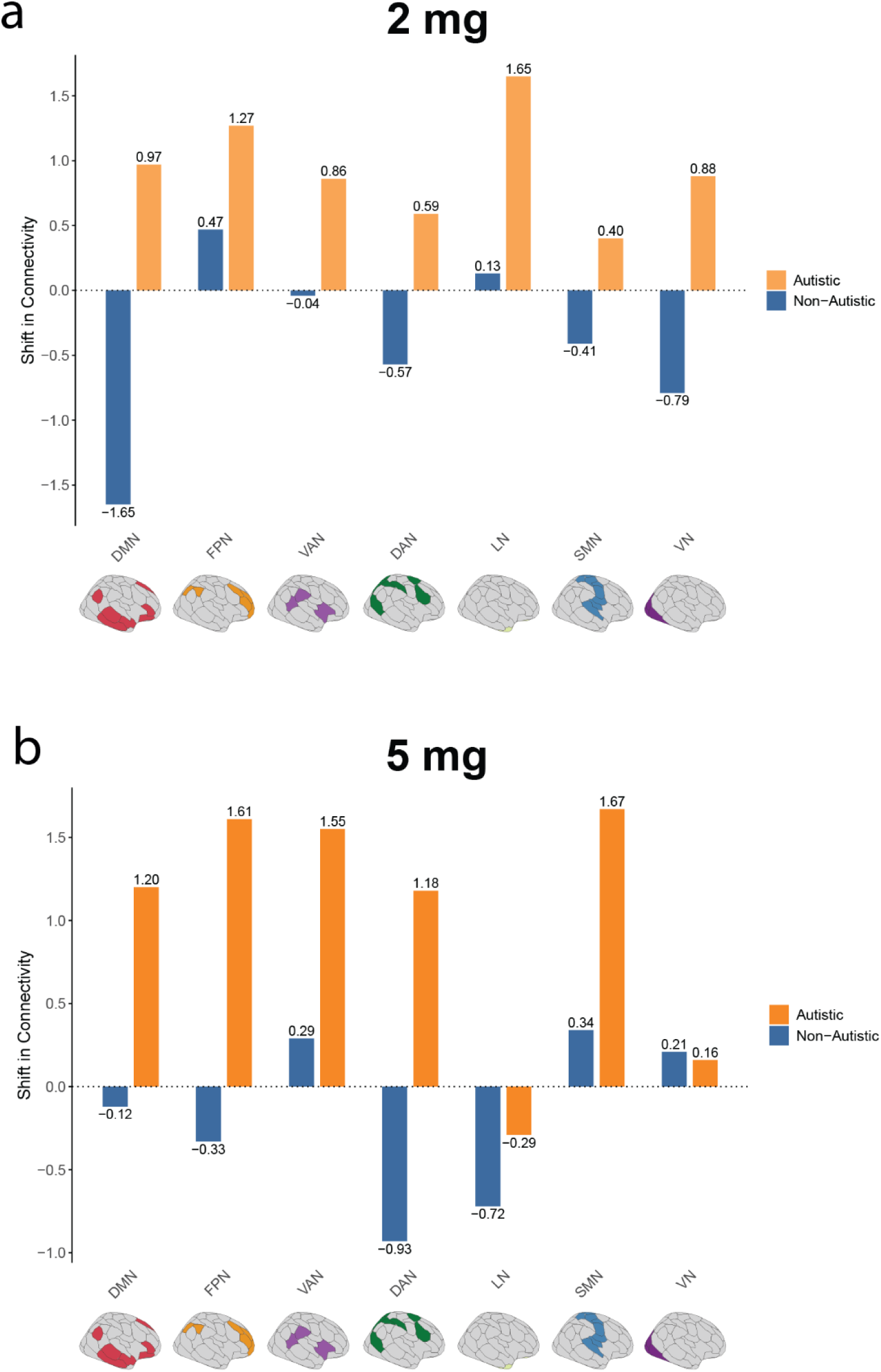
Total between-network connectivity shifts of each resting-state network in response to **(a)** 2 and **(b)** 5 mg psilocybin in non-autistic and autistic participants. Values represent the total between-network connectivity of a given network (visualised as an aggregate shift index computed as the sum of the edgewise effect sizes from the corresponding between-network matrices); at 5 mg psilocybin, increased overall between-network connectivity in the autistic group was driven by increased connectivity between several higher-order networks (e.g. DMN, FPN, VAN). DMN, default mode; FPN, frontoparietal; VAN, ventral attention; DAN, dorsal attention; LN, limbic; SMN, somatomotor & VN, visual network.

**Figure 5.**
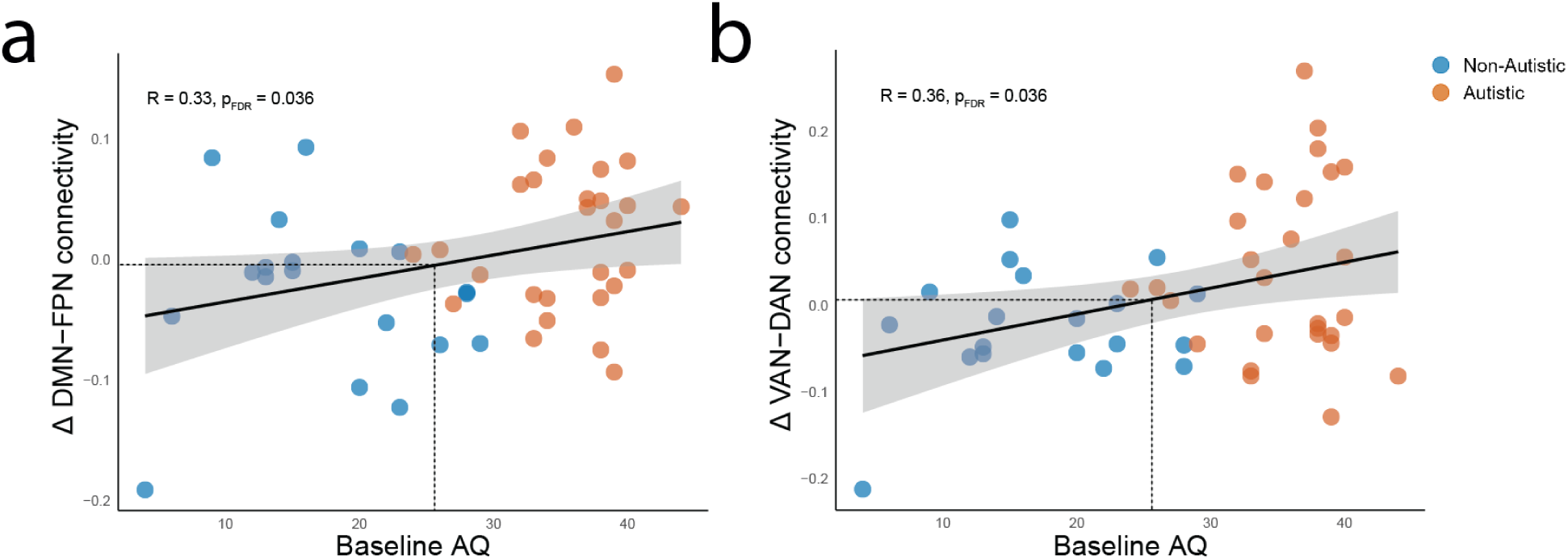
The relationship between change in **(a)** DMN-FPN and **(b)** VAN-DAN connectivity with baseline total Autism Quotient (AQ) scores showing increased between-network connectivity after 5 mg psilocybin is associated with higher core autistic traits at baseline across the whole cohort. The dotted line represents the clinical screening threshold for significant autistic traits of ≥ 26. Non-autistic and autistic participants are shown in blue and orange, respectively.

In summary, DMN-FPN and DAN-VAN connectivity was decreased by 5 mg psilocybin in non-autistic participants and increased in autistic participants. 5 mg psilocybin decreased within-network connectivity in non-autistic participants in the FPN and LN and increased FPN-SMN between-network connectivity in autistic participants. Globally, between-network connectivity was increased in autistic participants after both 2 and 5 mg psilocybin.

### Network connectivity alterations by psilocybin correlated with the extent of core autistic traits

Finally, significant shifts in network connectivity after 5 mg psilocybin were correlated with the extent of core autistic traits across the whole group (quantified using AQ total score). Reduced within FPN connectivity was correlated with lower AQ scores (β = 0.32, T = 2.14, p_FDR_ = 0.036) – consistent with the shift observed in non-autistic individuals. Increased between DMN-FPN (β = 0.33, T = 2.16, p_FDR_ = 0.036), DAN-VAN (β = 0.36, T = 2.43, p_FDR_ = 0.036) and FPN-SMN (β = 0.34, T = 2.18, p_FDR_ = 0.036) connectivity following 5 mg psilocybin was associated with higher AQ scores – consistent with the shift observed in autistic people.

### Subjective effects of low dose psilocybin in non-autistic and autistic people were modest and generally comparable

There was no significant main effect of group on 5D-ASC total score, thus non-autistic and autistic participants did not differ at baseline (i.e. placebo).

The 5 mg psilocybin dose significantly increased the subjective effects experienced by participants compared to placebo and their overall intensity was comparable in the non-autistic and autistic group **(Supplementary Figure 2)**.

Low doses used here did not induce *marked* psychedelic experiences in either group. The subjective effects elicited compared with a previous study of 3 and 6 mg psilocybin^11^. For context, high dose psilocybin (∼8-30 mg) results in subjective effect scores on average double those reported here at 5 mg^32,33^.

## Discussion

We report for the first time that low dose psilocybin differentially shifts resting-state network functional connectivity in autistic individuals.

In line with our hypotheses, the 5 mg psilocybin dose elicited the greatest shifts in functional connectivity in both groups, but in different directions. In general, 5 mg psilocybin decreased *within*-network connectivity in non-autistic participants and increased *between*-network connectivity in autistic participants. Across the whole cohort, these network shifts were correlated with autistic traits – those with least reduction in within-network connectivity and largest increase in between-network connectivity in response to 5 mg psilocybin had the most autistic traits.

Despite this differential response of resting-state network connectivity after psilocybin in each group, the subjective effects reported by participants were comparable. Thus, what distinguishes the response of autistic and non-autistic individuals to psilocybin is not subjective experience, but objective neural responses and these relate to the extent of autistic traits as measured on the AQ.

### Non-autistic response to low dose psilocybin

In non-autistic individuals, a general decrease in within-network connectivity elicited by psilocybin in healthy volunteers is a reproducible finding^19^. In this study, 2 mg psilocybin elicited minimal shifts in connectivity, but 5 mg psilocybin decreased FPN and LN connectivity, which is in line with previous reports^7,34^. Thus, as expected, the network connectivity response to low dose psilocybin was in the same direction but to a lesser extent than higher doses in non-autistic people.

This may, in part, be explained by a lower occupancy of the 5HT_2A_ receptor at lower doses of psilocybin^11^. 5HT_2A_ receptor occupancy has a fundamental role in the high dose response^35^ - 5HT_2A_ receptor occupancy by psilocin (the active metabolite) on the layer V pyramidal neurons is responsible for the cross-cortical organisation of neuronal firing rates^36^, which causes desynchronisation of brain activity within-networks to increase global connectivity and that of less functionally connected networks i.e. between-network connectivity^5–8^. The degree of cortical excitation induced by 5HT_2A_ receptor signalling at low doses may not be sufficient to act between-networks, so the effect is limited to brain regions with higher coherence at rest i.e. within-networks. That is, in non-autistics, the threshold to shift the brain out of its equilibrium state may be lower within-networks, but starts to occur only at 5 mg.

Psilocybin has complex pharmacology and binds several other serotonin receptors^37,38^ which may also be involved. Psilocybin has higher affinity for the 5HT_1A_ receptor than 5HT_2A_ receptor^37^ and at low dose there may be a bias towards 5HT_1A_ agonism. This may particularly contribute to the decreased limbic connectivity in non-autistic participants observed here, comprising regions rich in 5HT_1A_ receptors^39^.

### Differential autistic response to low dose psilocybin

In contrast, low doses of psilocybin were sufficient to increase between-network connectivity in autistic people. This distinct effect cannot be easily explained by baseline group differences as brain connectivity did not differ in the placebo condition. Instead, they are *responsivity* differences. Possible explanations are that the lower 5HT_2A_ receptor levels and fewer parvalbumin-expressing interneurons (which also express 5HT_2A_ receptors^40^), which have been reported in the autistic brain^41^, shape the response to psilocybin. These interneurons typically synchronise the neuronal activity of pyramidal neurons via GABA release following 5HT_2A_ receptor activation and constrain neural activity temporally and spatially. If this does not happen, then elevated connectivity between functionally segregated networks may increase, as observed here in autistic participants. Such a mechanism would fit with evidence for interneuronal and GABAergic differences in autism^41^.

Another, or additional, explanation may be that network connectivity changes, especially at single, acute low doses with a shorter duration of action, may persist longer in autistic people and are captured at the timepoint of data collection. This homeostatic account is supported by preclinical models relevant to autism^42^. It also fits with our previous work in which we reported increased between-network connectivity^10^ (effectively greater network integration) in response to drug challenge across multiple neurotransmitter systems in autism.

Another parsimonious explanation is simply that autistic people are more ‘sensitive’ to psilocybin as low dose effects mirror those reported at higher doses in assumed non-autistic people^19^. This may have translational implications because the therapeutic effects of psilocybin rely on increased global brain integration^9^, including between higher-order association networks (i.e. DMN-FPN)^43^ as observed at low dose here.

An implication of our study is that lower doses of psilocybin may potentially be sufficient for treatment of depression and related mental health difficulties in autism. Our study cannot speak to this directly but this warrants further investigation. There would be pragmatic benefits. In clinical trials, ‘psilocybin treatment’ with a high dose of psilocybin that induces a marked psychedelic experience requires a psychological support model (^44^for example; eye shades and headphones, trained therapists etc.). However, this paradigm may not necessarily be suitable for autistic people and particularly those with intellectual disabilities and non-speaking individuals. There is still a long way to go to understand how best to adapt existing psychotherapies in autistic people, doing so in the context of a marked psychedelic experience adds further complexity. Thus, low dose psilocybin may be a more tractable way forward.

Another future direction would be to harness the *individual* responses to psilocybin found here to improve clinical outcomes for autistic people. For example, increased between-network connectivity after psilocybin has recently been proposed as a ‘biomarker’ of depression response^45^. Here, this increase was associated with the extent of core autistic traits at baseline. This suggests that the underlying functional differences in the serotonergic targets of psilocybin exist along a continuum within the general population, like autistic traits themselves^17^, as opposed to binary group differences in response based on behavioural diagnosis of autism alone. Thus, our findings may help address heterogeneity and better personalise future interventions. Together with improved pre-trial measures of target engagement, this will inform a more personalised approach to psilocybin treatment. An important next step would also be to examine whether there are autistic differences in the post-treatment connectivity alterations associated with clinical response^43^.

There are several limitations to our work. First, the fMRI methods applied here were intended as an initial exploratory approach and used a well-validated parcellation scheme in the psychedelic and autism literature^9,34,46,47^. However, whole-brain and seed-based approaches that include sub-cortical regions, for example, which are important in psilocybin drug action^48,49^ will be a next step. Second, our study lacks pharmacokinetic data (such as plasma psilocin levels). Plasma psilocin is known to influence both subjective effects and network changes in response to psilocybin^7,11^, and so group differences in psilocybin metabolism may have influenced the connectivity metrics reported here as the scan at peak effects (∼60 mins) may not reflect the peak in autistic individuals. Further, polymorphisms in genes associated with drug metabolism (e.g. CYP2D6, which is involved in the metabolism of psilocybin^50^) have previously been associated with variability in safety and side effects of risperidone treatment in autistic individuals, for example^51^. Whether the pharmacokinetics of psilocybin are altered in autism, and impacted our results, remains unknown.

In summary, the response of resting-state network connectivity to low dose psilocybin is different in non-autistic and autistic people and related to the extent of autistic traits, despite comparable subjective effects. We suggest that future applications of psilocybin in autistic people should consider the utility of lower doses. Clinical trials for mental health conditions do not typically screen for autism despite the likelihood that this population is over-represented in mental health settings and often report greater sensitivity to low doses of psychoactive medication, more treatment-resistance, more side effects and paradoxical responses^4^. Thus, an autistic brain that responds differently to psilocybin will be crucial to recognise to ensure safety and equity of access.

## Data Availability

Data may be made available on reasonable request, subject to data access agreements with the study sponsor and other rights holders, and to any conditions imposed by the relevant research ethics committee.

## Supplementary Information

**Supplementary Figure 1.**
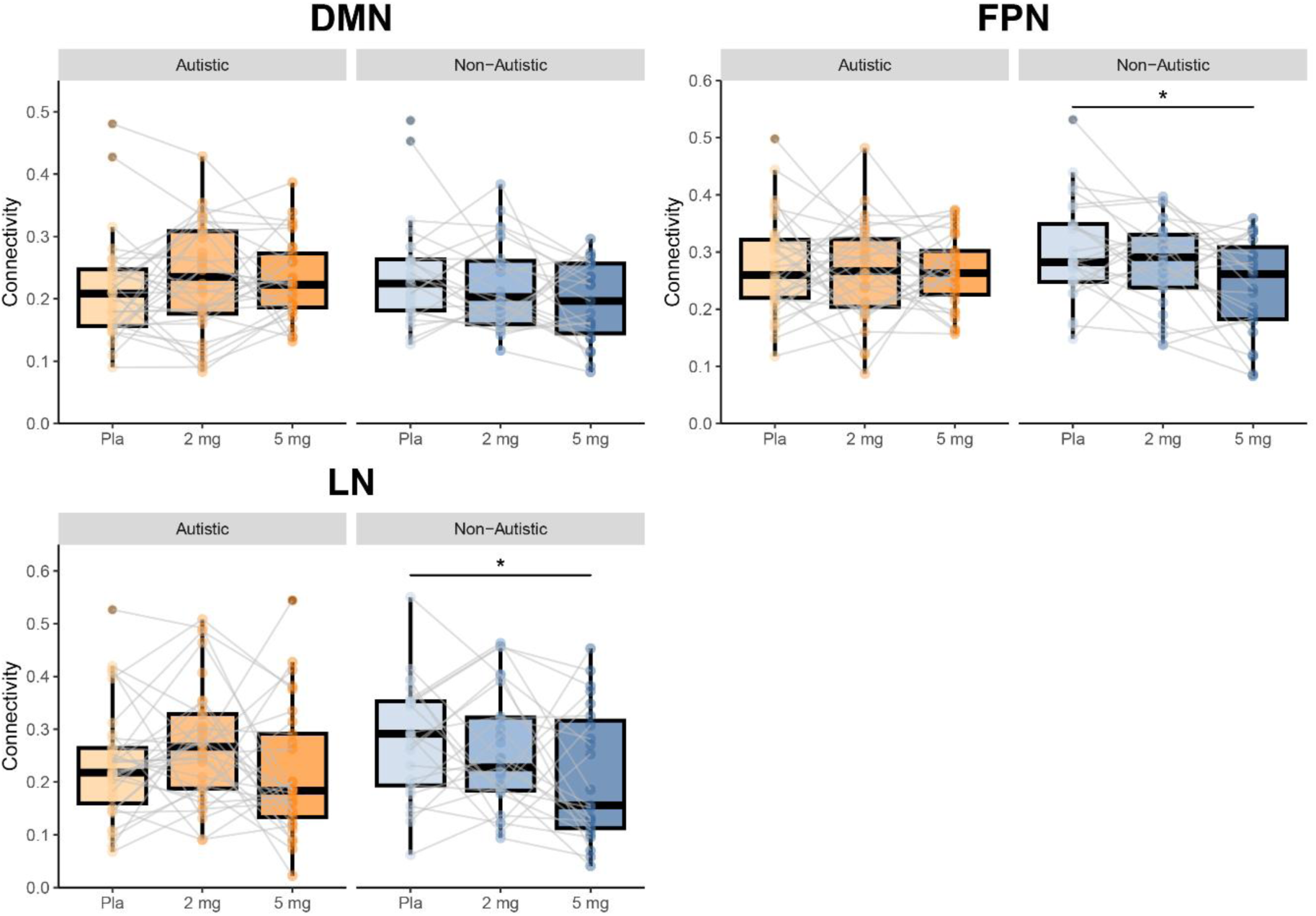
Within-network connectivity alterations in response to psilocybin in non-autistic and autistic participants. There was a significant dose × group interaction for the DMN but this did not survive correction for multiple comparisons. There was a main effect of dose for the FPN and LN, post-hoc analyses revealed that this was driven by reductions in FPN and LN connectivity after 5 mg in the non-autistic group (*p_FDR_ < 0.05). DMN, default mode; FPN, frontoparietal & LN, limbic networks.

**Supplementary Figure 2.**
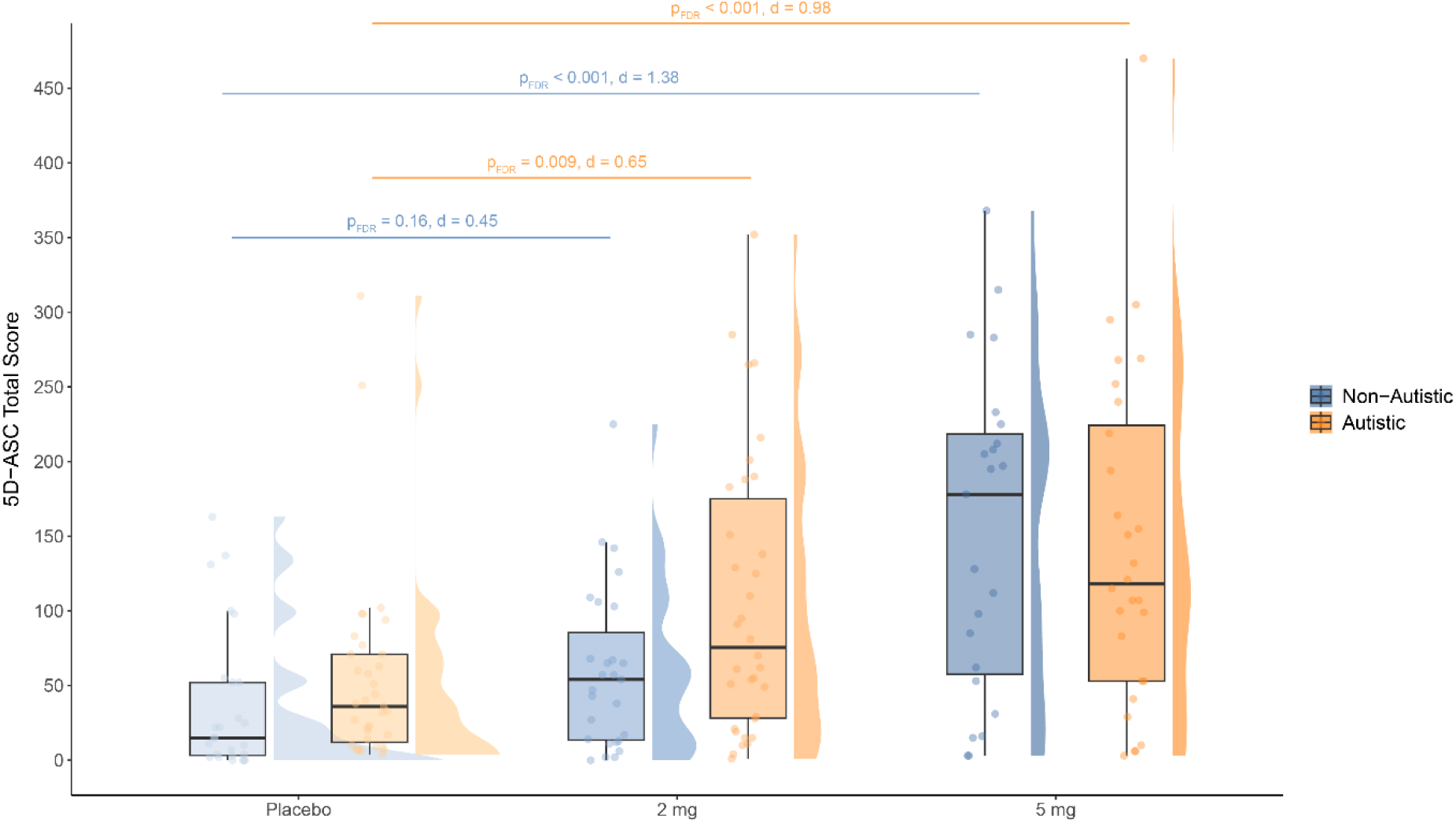
5D-ASC total scores in non-autistic and autistic individuals. Significant within-group main effect of dose comparisons with placebo are represented with a dagger (p_FDR_ < 0.05). ns, not statistically significant. There was no significant dose × group interaction for 5 mg on 5D-ASC total score (β = −23.9, T = −0.90, p_FDR_ = 0.37). There was a significant main effect of dose at 5 mg psilocybin (β = 118.6, T = 6.01, p_unc_ < 0.001). Post-hoc analyses confirmed a main effect of dose at 5 mg in both non-autistic (β = 118.6, T = 5.99, p_unc_ < 0.001) and autistic participants (β = 95.2, T = 5.48, p_unc_ < 0.001). There was also no significant dose × group interaction for 2 mg on 5D-ASC total score (β = 26.2, T = 1.04, p_FDR_ = 0.30). Post-hoc analyses revealed that the 2 mg psilocybin dose significantly increased 5D-ASC total score compared to placebo in autistic (β = 53.6, T = 3.26, p_FDR_ = 0.009, d = 0.65), but not non-autistic participants (β = 28.3, T = 1.44, p_FDR_ = 0.16, d = 0.45). 5D ASC, Five-Dimensional Altered States of Consciousness.

**Supplementary Figure 3.**
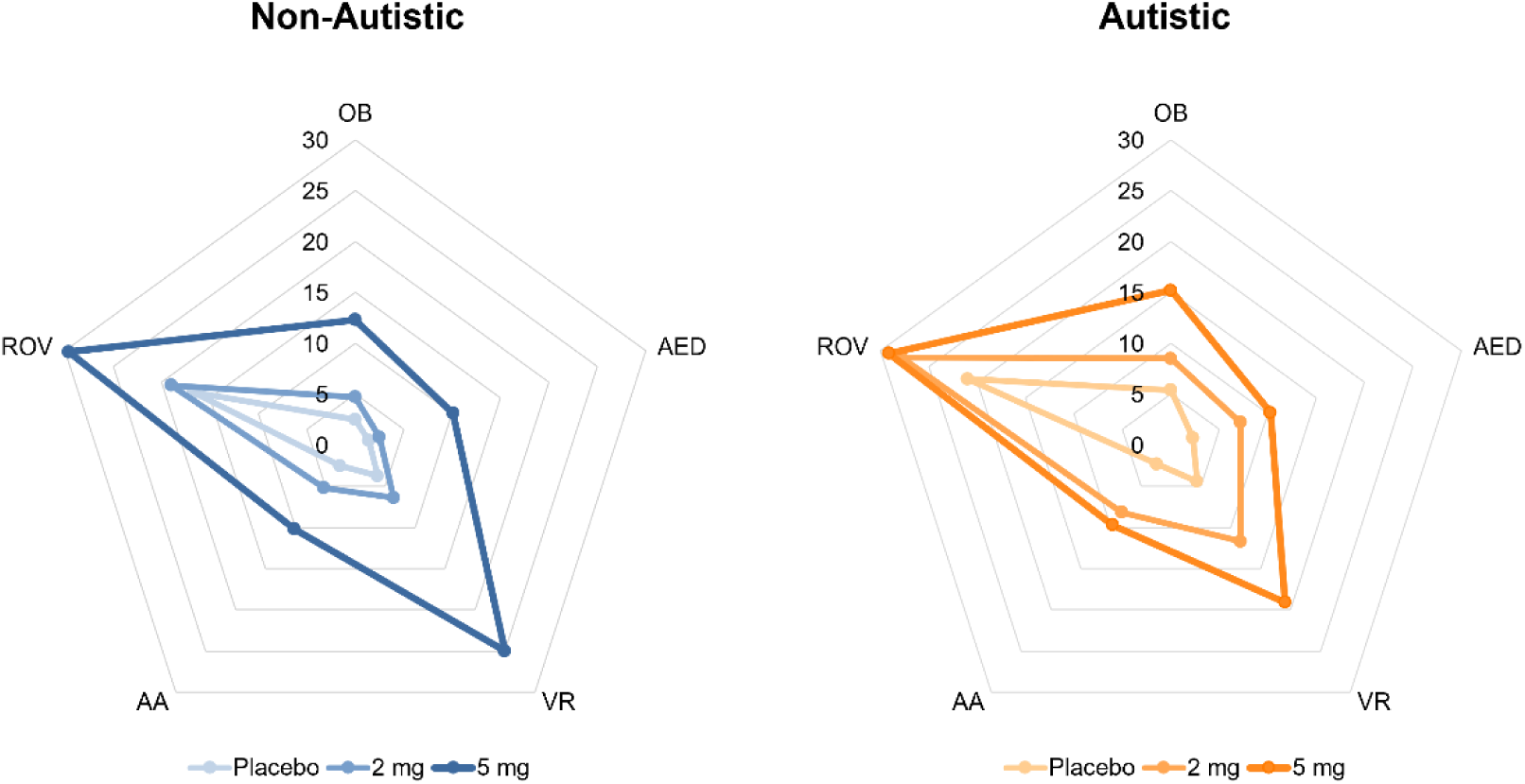
5D-ASC dimension scores for non-autistic and autistic participants in response to placebo, 2 mg and 5 mg of psilocybin. Values represent the percentage of maximum total score for each dimension. 5D-ASC dimension scores were no different at baseline and all dimension scores increased after 5 mg psilocybin; autistic and non-autistic participants also did not differ on dimensional response to psilocybin **(Supplementary Table 2** for full 5D-ASC dimensional results after 5 mg psilocybin). OB, oceanic boundlessness; AED, anxious ego dissolution; VR, visionary restructuralisation; AA, auditory alterations & ROV, reduction of vigilance.

---

**Supplementary Table 1.**
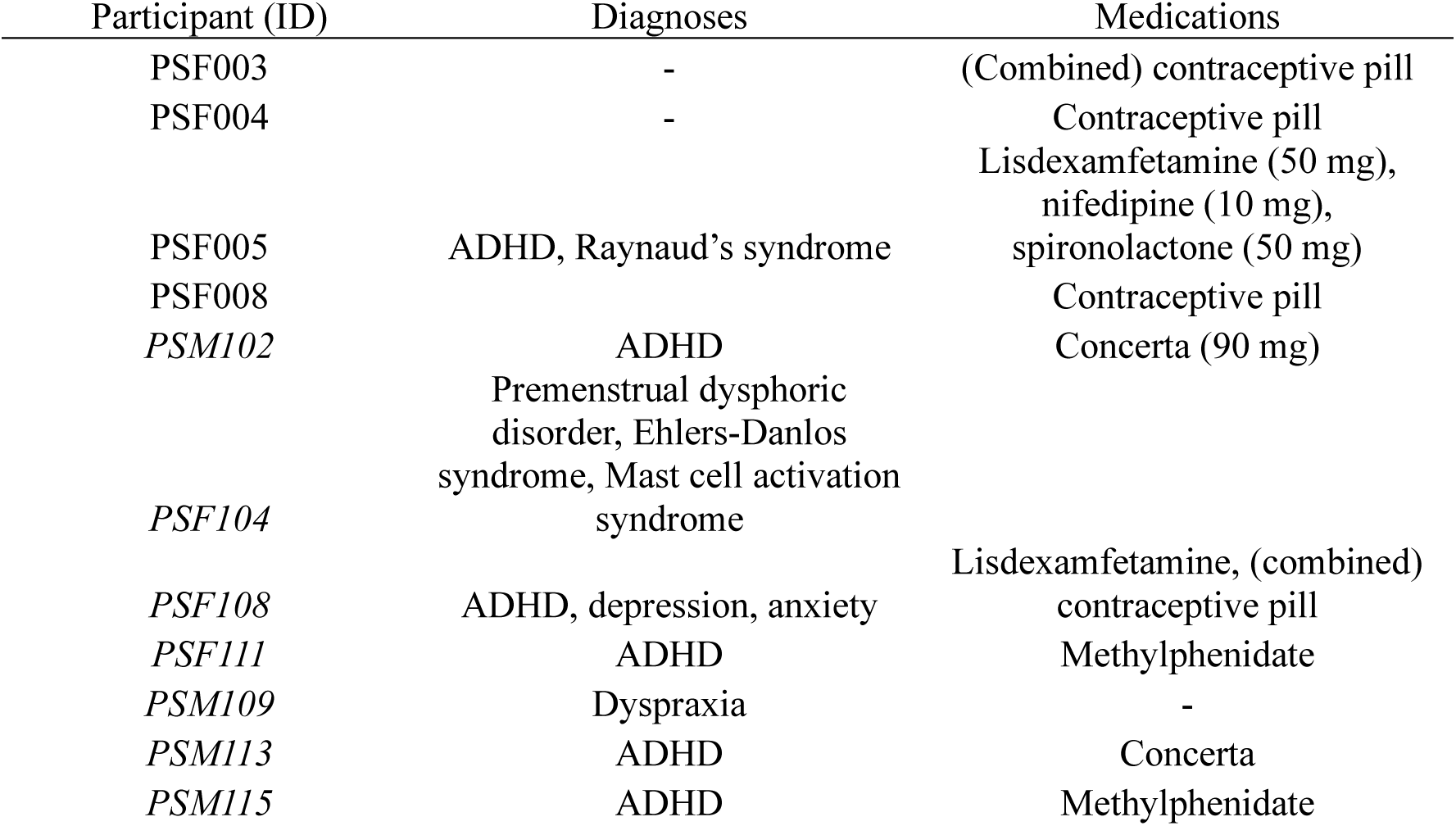
Further information on concurrent medications taken by ‘PSILAUT’ study participants. M or F in participant ID denotes male or female, respectively; autistic participants are shown in italics.

**Supplementary Table 2.** The within-group main effect of dose for dimensions scores of the 5D-ASC questionnaire in non-autistic individuals. Dose represents the pairwise comparison between the placebo and 5 mg psilocybin dose level. P values were corrected using the FDR method across dose levels for each dimension separately. Effects sizes are represented by Cohen’s d. OB, oceanic boundlessness; AED, anxious ego dissolution; VR, visionary restructuralisation; AA, auditory alterations and ROV, reduction of vigilance.

|  | Dose | Non-Autistic |  |  |  |  | Autistic |  |  |  |  |
| --- | --- | --- | --- | --- | --- | --- | --- | --- | --- | --- | --- |
| | | $\beta$ | T | $P_{unc}$ | $P_{FDR}$ | d | $\beta$ | T | $P_{unc}$ | $P_{FDR}$ | d |
| OB | 0, 5 | 1.01 | 3.88 | <0.001 | <0.001 | 1.15 | 1.14 | 4.95 | <0.001 | <0.001 | 0.76 |
| AED | 0, 5 | 0.89 | 4.16 | <0.001 | <0.001 | 1.05 | 0.85 | 4.52 | <0.001 | <0.001 | 0.77 |
| VR | 0, 5 | 2.15 | 6.60 | <0.001 | <0.001 | 1.43 | 1.48 | 5.14 | <0.001 | <0.001 | 1.13 |
| AA | 0, 5 | 0.80 | 3.24 | 0.001 | 0.003 | 0.78 | 0.83 | 3.81 | <0.001 | <0.001 | 0.75 |
| RoV | 0, 5 | 1.35 | 3.04 | 0.002 | 0.006 | 0.66 | 0.88 | 2.25 | 0.026 | 0.026 | 0.40 |

## Acknowledgements

Competing Interests: T.P.W. and E.M. are co-founders and hold shares in N1 Bio Corp. and hold shares in Compass Pathfinder Ltd. N.A.P. has consulted for Deerfield Discovery and Research. D.G.M.M. has consulted for Jaguar Gene Therapy LLC and N1 Bio Corp. G.M.M. has received funding for investigator-initiated studies from GW Pharmaceuticals and Compass Pathfinder Ltd. G.M.M. has consulted for Greenwich Biosciences, Inc. and N1 Bio Corp. M.D., L.G.S.F., C.L.E., F.M., F.M.P., J.K., N.K., Y.G., N.M., G.I., E.D., and D.B. have no conflicts of interest to declare.

## Funding

The study was an investigator-initiated study part funded by Compass Pathfinder Ltd. The authors also receive support from EU-AIMS (European Autism Interventions)/EU AIMS-2-TRIALS, an Innovative Medicines Initiative Joint Undertaking under Grant Agreement No. 777394. In addition, this paper represents independent research part funded by the infrastructure of the National Institute for Health Research (NIHR) Biomedical Research Centre (BRC): Maudsley and the Medical Research Council Centre (MRC) for Neurodevelopmental Disorders. The views expressed are those of the author(s) and not necessarily those of the NHS, the NIHR or the Department of Health and Social Care.

## Notes

### Clinical Trial

NCT05651126

### Clinical Protocols

https://link.springer.com/article/10.1186/s12888-024-05768-2

### Author Declarations

Written informed consent was obtained from all participants in accordance with the Helsinki Declaration of 1964, as revised in 2013 and the study procedures were approved by Dulwich Research Ethics Committee (21/LO/0795).

